# The local stability of a modified multi-strain SIR model for emerging viral strains

**DOI:** 10.1101/2020.03.19.20039198

**Authors:** Miguel Fudolig, Reka Howard

**Affiliations:** Department of Statistics, University of Nebraska-Lincoln, Lincoln, NE, USA

## Abstract

We study a novel multi-strain SIR epidemic model with selective immunity by vaccination. A newer strain is made to emerge in the population when a preexisting strain has reached equilbrium. We assume that this newer strain does not exhibit cross-immunity with the original strain, hence those who are vaccinated and recovered from the original strain become susceptible to the newer strain. Recent events involving the COVID-19 virus demonstrates that it is possible for a viral strain to emerge from a population at a time when the influenza virus, a well-known virus with a vaccine readily available for some of its strains, is active in a population. We solved for four different equilibrium points and investigated the conditions for existence and local stability. The reproduction number was also determined for the epidemiological model and found to be consistent with the local stability condition for the disease-free equilibrium.

## Introduction

More recently, the anti-vaccination movement has been gaining traction in different parts of the world. Individuals who do not advocate vaccination commonly cite reasons such as fear of adverse side-effects, perceived low efficacy of vaccines, and perceived low susceptibility to diseases amongst others [1–3]. The drop in numbers in vaccination has led to outbreaks of diseases such as mumps [4–6] that could have been prevented by vaccination. Another example would be measles, which was declared eliminated in the United States back in 2000, has had outbreaks reported in the country since 2008 [10]. According to the Centers for Disease Control and Prevention (CDC), the reemergence is due to the presence of unvaccinated individuals and their interaction with other people who got the disease from other countries such as Israel, Philippines, and Micronesia [8–10]. According to the CDC, 1,282 individual cases of measles have been confirmed in 31 states in 2019, the highest since 1994. [10]

Another way for a disease to reemerge is through change in its antigenic properties, which is the case for the influenza virus. The influenza virus can mutate in two ways: through antigenic shift or antigenic drift [11–13]. Antigenic drift is defined as the result of frequent mutations of the virus, which happens every 2-8 years. On the other hand, the antigenic shift occurs around thrice every one hundred years and only happens with influenza A viruses [13]. Although more unlikely to happen than the antigenic drift, the antigenic shift involves genetic reassortment which can make it feasible to create a more virulent strain than the original strain [13–15].

One way to model the dynamics of an infectious disease is through compartmental systems. This approach involves separating the population into multiple components and describing infection and recovery as transitions between the set components. The simplest compartmental model to describe a viral infection is called the standard Susceptible-Infected-Removed (SIR) model, the dynamics of which has been studied in different references [16–19]. The SIR model separates the population into three compartments: the susceptible (S), infected (I), and removed (R) compartments. The susceptible compartment is comprised of individuals that are healthy but can contract the disease. The infected compartment is comprised of individuals who have already contracted the disease. Lastly, the removed compartment is comprised of individuals who have recovered from the disease. Individuals who have recovered from a certain strain of a viral infection are likely to be immune to infection of the same strain [20, 21], which is why the SIR model is used to model viral infections. SIR models can provide insight on the dynamics of the system and has been used to model different influenza virus strains such as the swine and avian flu focusing on the spatio-temporal evolution and equilibrium dynamics of the system for both disease free and endemic equilibrium cases [22–27].

One important parameter resulting from the SIR models is the reproduction number. The reproduction number of an infectious disease is defined as the expected number of secondary infections caused by a single infected individual for the whole duration that they are infectious [28, 29]. The reproduction number *R*_0_ describes how infectious a disease can be, and can also be used as a threshold parameter to determine whether a disease would survive in a healthy population [29].

Modifications of the SIR model have been used to model mutations and changes in an infectious virus such as influenza (flu). Yaari et al. [30] used a discrete time stochastic susceptible-infected-removed-susceptible (SIRS) model to describe influenza-like illness in Israel accounting for weather and antigenic drift by adding terms that account for weather signals and lost of immunity. Finkenstadt et al. [31] created a predictive stochastic SIRS model for weekly flu incidence accounting for antigenic drift. Roche et al. [32] used an agent-based SIR model to describe the spread of a multi-strain epidemic, while Shi et al. [33] used the same approach and empirical data from Georgia, USA to model an influenza pandemic that incorporates viral mutation and seasonality. However, these approaches have been stochastic in nature, which does not provide information regarding the stability and existence of equilibrium points in an infected population. The abovementioned articles also do not take vaccination and the presence of other strains into account in their models. Consequently, one can model the presence of a mutated virus spreading into a population using a multi-strain model, which was used in the following studies for avian flu [23, 24]. These models study the birds and the humans as one population in an SI-SIR model, where the first SI corresponds to the compartments for the avian species. However, the two infected compartments in this model do not cross since they are separated by species. Casagrandi et al. [25] introduced a non-linear deterministic SIRC epidemic model to model the influenza A virus undergoing an antigenic drift. The SIRC model is a modified SIR model with an additional compartment, C, for individuals that receive partial immunity from being infected by one of the present strains. Although able to account for cross-immunity between strains, the model does not include the effect of vaccination into the system. Papers which have considered vaccination only consider one strain propagating within the population [34–37]. There has been very few studies that investigate the effect of vaccination in the presence of multiple strains like Wilson et al. did for Hepatitis B [38], which did not investigate the equilibrium model in detail.

In the case of the influenza virus, it is possible to have multiple strains exist in a population, but only have vaccine for a certain strain that will not be effective for others. The fact that viruses undergo changes regularly indicates that people who have recovered from the virus, as well as individuals who have been vaccinated for a specific strain of the virus, can be susceptible again to a newly-emerged strain. It is important to determine the conditions in which a newly emerged strain and a common strain that has a means of immunity will coexist in a population.

Another apt example for emerging diseases is the emergence of the COVID-19 virus in 2019 [39, 40]. As of March 17, 2020, there have been 167,545 confirmed cases of COVID-19 in 150 different countries that has led to 6,606 deaths since it was declared as an outbreak in January 2020 according to the WHO situation report [41]. At the time that this paper is being written, there are papers that have modeled the dynamics of the virus using different modifications of the SIR model. Zhou et. al. [42] included compartments corresponding to suspected cases, which consists of the individuals that show similar symptoms but are not confirmed cases, and indirectly infected individuals. Pan et. al. [43] used a modified SEIR model which included asymptomatic and treatment compartments for occurrences in Wuhan, China, the city where the outbreak started, and outside of Wuhan. Maier and Brockmann [44] included a separate compartment for quarantined individuals in the SIR model to account for the containment measures applied by the public for the virus. They then estimated the reproduction number of COVID-19 in different locations in China. It is notable that this virus emerged during the flu season [45] and had managed to infect a lot of individuals around the world in such a short time even when the threat of the influenza virus still exists. This paper will give researchers insight about the conditions in which one strain can dominate another or if two different strains can coexist in a population, given that one of these strains has a vaccine available. This paper introduces a model that approaches the lack of cross-immunity across different viral strains by introducing new compartments to the SIR with vaccination model. This enables us to introduce acquired immunity through vaccination and cross-immunity between strains in a simple compartmental model and investigate the existence and stability of the resulting equilibrium points.

We aim to model the dynamics of an epidemic where a new emergent strain of an existing virus affects a closed population. The existing virus will be modeled using a modified SIR model with vaccination, however we assume that the vaccine does not provide immunity to the newer strain. The equilibrium points were determined for the system based on the transition equations and local stability was investigated for each point. Once the stability conditions have been established, the epidemic model was simulated using R [46] to check the steady-state behavior of the surveillance data for each compartment of the population. The values for the transmission and removal coefficients were dictated by the existence and stability conditions for each equilibrium point during the simulation. The reproduction number for the epidemic was also determined for this modified SIR epidemic model and compared to existing SIR models.

## Modelling the emergence of the new strain

This section describes how the emergence of the new strain of the virus will be incorporated into the model. This emergence can either be due to mutation, antigenic drift/shift, or an introduction of a different strain from an external source. Assume that initially, there is only one strain of the virus that exists in the population. Immunity can be achieved either by recovering from the infection or getting vaccinated. After equilibrium has been established with the original strain, the new strain is introduced to the population. The new strain can affect individuals previously infected by the original strain and those who are vaccinated against the original strain; the only way to be immune to the mutated strain is to recover from the infection of the new strain. The next two subsections will explain the dynamics before and after the emergence of the mutated strain.

### Before emergence

The system starts off as a population exposed to the original strain of the virus. The spread of the virus is described by a modified SIR model that accounts for vaccination [34]. The vaccinated members of the population can be treated as members of an additional compartment that do not interact with the infected individuals. This means that the modified SIR model will have four compartments instead of three, which are given by:

- Susceptible **S**: Individuals in this compartment are healthy, but are susceptible to be infected by the disease since they are not vaccinated.
- Vaccinated **V**: Individuals that were given a vaccine, making them immune to the disease. This also includes individuals with natural immunity to the disease.
- Infected **I**_1_: Individuals that are infected by the disease.
- Removed **R**: Individuals that were infected but are now immune to the disease upon recovery. Because of their immunity, the members of this compartment do not interact with the remaining compartments.

Let *S, V, I*_1_, and *R* be the respective number of individuals in the susceptible, vaccinated, infected, and removed compartments. The transition between the compartments is summarized by the compartmental diagram shown in Fig 1.

**Fig 1.**
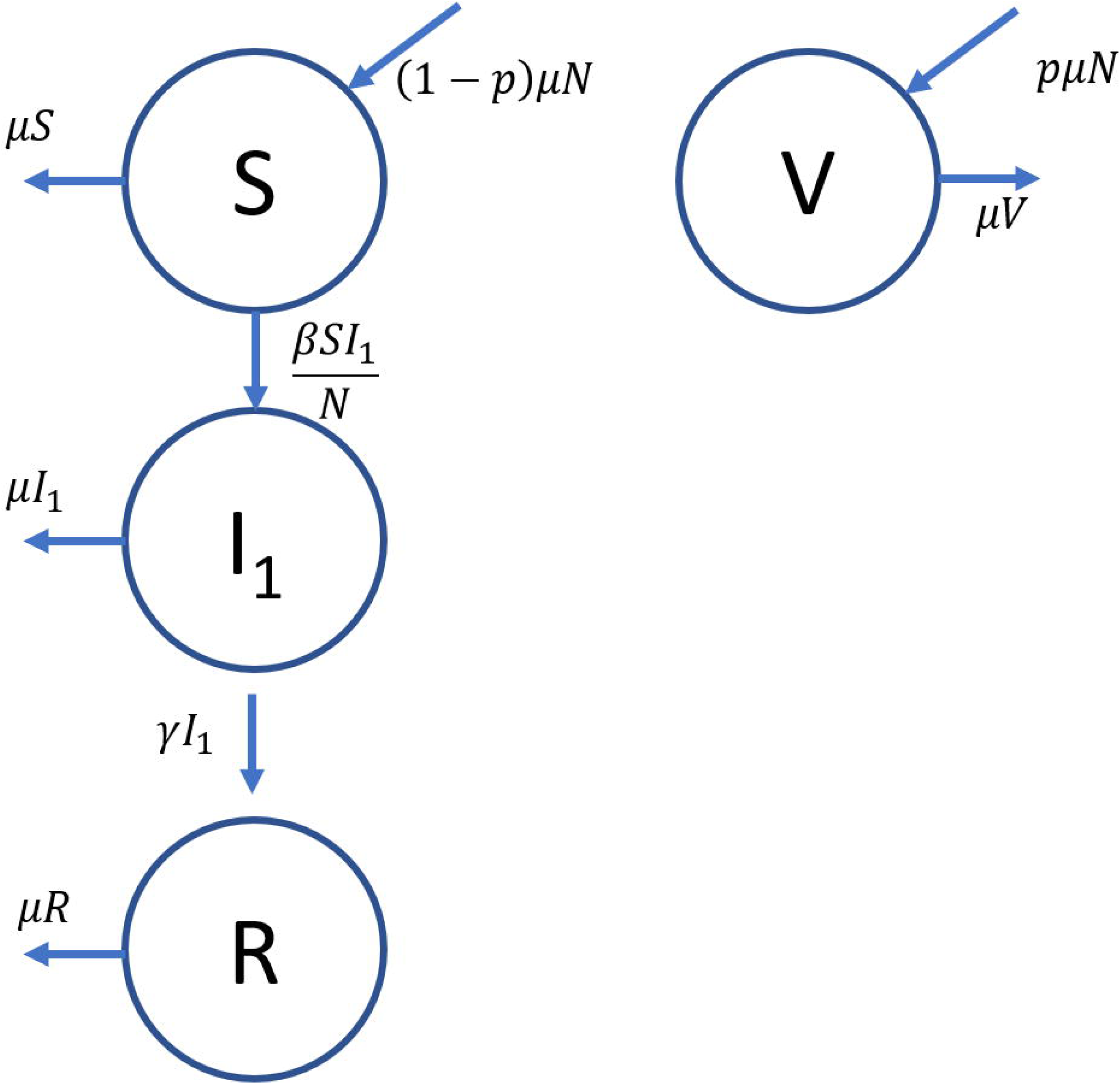
Compartment diagram with transitions for the SIR with vaccination model. The arrows show the transitions between the compartments, as well as the exits due to natural death. The transition rates are shown next to the arrows.

For this model, *μ* be the natural birth rate of the population, and consequently the natural death rate of the population to keep the population size constant. It is assumed that the individuals are vaccinated at birth with a vaccination rate *p. β* is the standard incidence transmission coefficient, which assumes that the infection occurs based on how many susceptible individuals interact with the infected [47]. For standard incidence, the contact rate between infected and susceptible individuals is constant over all infected individuals regardless of the population size [48]. The removal rate coefficient for the infected individuals is denoted by *γ*.

The dynamics of the system is described by ordinary differential equations that describe the rate of change in individuals belonging to a specific compartment. For any number of individuals in a general compartment *C*, the rate of change of membership in the compartment can be expressed by the following equation:

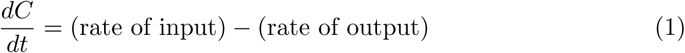

where the rates are denoted in the compartment diagram shown in Fig 1. For the susceptible compartment *S*, the rate of increase comes from the birth of new members of the population who are not vaccinated, which is given by (1 − *p*)*μN*. Meanwhile, susceptible individuals can either get infected at a rate of 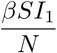 or die due to natural causes at a rate *μS*. In equation form, this translates to

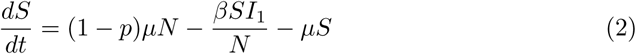

For the infected compartment *I*_1_, the number of infected individuals increase when susceptible individuals get infected at a rate 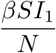. The infected individuals can either die at a rate of *μI*_1_ or get removed and not interact with the system again at a rate *γI*_1_ when they recover. Translating this to an ordinary differential equation yields

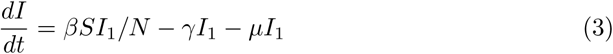

Unlike the members of the susceptible compartment, the individuals in the vaccinated compartment will not get infected by the virus. This implies that the changes in the number of vaccinated individuals can only be due to the rate of vaccination in the population and death due to natural causes. Using a similar approach, the rate equation for the vaccinated compartment *V* is given by

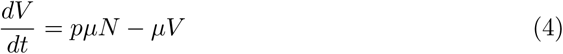

Since the population is closed, the number of individuals in the removed compartment can be expressed as *R* = *N* − *S* − *I*_1_ − *V*. This implies that solving Eqs 2–4 is enough to describe the system completely at any time *t*. Without loss of generality, Eqs 2–4 can be normalized with respect to the total population, *N*, so that the equations would be invariant to population scaling. This yields the following equations

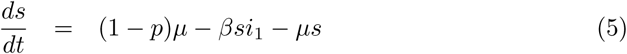

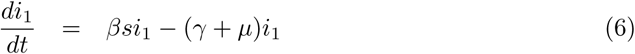

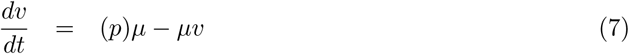

where (*s, i*_1_, *v*) = (*S/N, I*_1_*/N, V/N*) and *r* = *R/N* = *N* (1 − *s* − *i*_1_ − *v*) are functions of time *t*. Note that plausible solutions only exist when *s*(*t*), *i*(*t*), *v*(*t*), *r*(*t*) ≥ 0.

To achieve equilibrium, there should not be any changes in the proportion for each compartment, which implies that Eqs 5-7 should be zero. If Eq 6 is zero, then two conditions emerge: *i*_1_ = 0 or *i*_1_ =*/* 0. The first case corresponds to the disease-free equilibrium (DFE) point (*s*(*t*), *i*_1_(*t*), *v*(*t*)) = (1 − *p*, 0, *p*). The latter case corresponds to the endemic equilibrium point (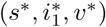). If *i*_1_ ≠ 0, then the following condition should be satisfied

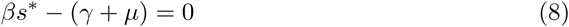

Solving for *s*^*^, we get

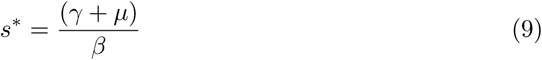

We can use this result to solve for 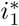 in Eq 6. The resulting endemic equilibrium point (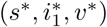) is given by,

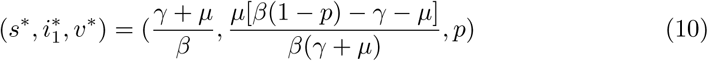

According to Chauhan et. al [34], the reproduction number of the disease for the vaccinated SIR model is given by *R*_*v*_ = *R*_0_(1 − *p*) = *β*(1 − *p*)*/*(*γ* + *μ*). This means that the endemic equilibrium point will be asymptotically stable if *R*_*v*_ > 1, while the DFE will be asymptotically stable if *R*_*v*_ < 1. The reproduction number will be discussed in the section “Reproduction Number”.

### Emergence of new strain

Suppose that at time *T* when equilibrium has been achieved in the SIR with vaccination model, a new strain of the disease is introduced to the population. This new strain will have a different transmission coefficient *β′* and removal rate coefficient *γ′*. This results to the existence of another compartment **I**_2_ for those who are infected with the new strain, which we will refer to as Disease 2.

The existence of the newer strain will be constrained by the following assumptions:

- Since the vaccine is assumed to only work on the original strain, the vaccinated and the previously removed individuals are susceptible to the newer strain.
- Once infected by the newer strain, the individual cannot be infected by the original strain. The individuals infected by the newer strain will be removed from the population or die.
- Individuals infected by the original disease have to be removed first before being susceptible to the newer strain; meaning that there is no chance of super-infection (**I**_1_ → **I**_2_) [28].

This means that the number of compartments that need to be monitored will increase from four to six, with the addition or modification of the following compartments:

- **R**_1_: Individuals who have recovered from the original strain but are now susceptible to the newer strain.
- **I**_2_: Individuals who are infected by the newer strain.
- **R**_2_: Individuals who were previously infected by the newer strain but have now been removed due to recovery or treatment.

The members of the vaccinated compartment, which was initially an isolated compartment, can now be infected by the new strain. The same can be said for the individuals who have recovered from the original strain. For mathematical simplicity, the infection coefficients for the new strain are assumed to be the same for the susceptible, vaccinated, and initially recovered compartments.

These assumptions and the increase in number of compartments also introduce the possibility of new transitions between compartments as shown in Fig 2:

**Fig 2.**
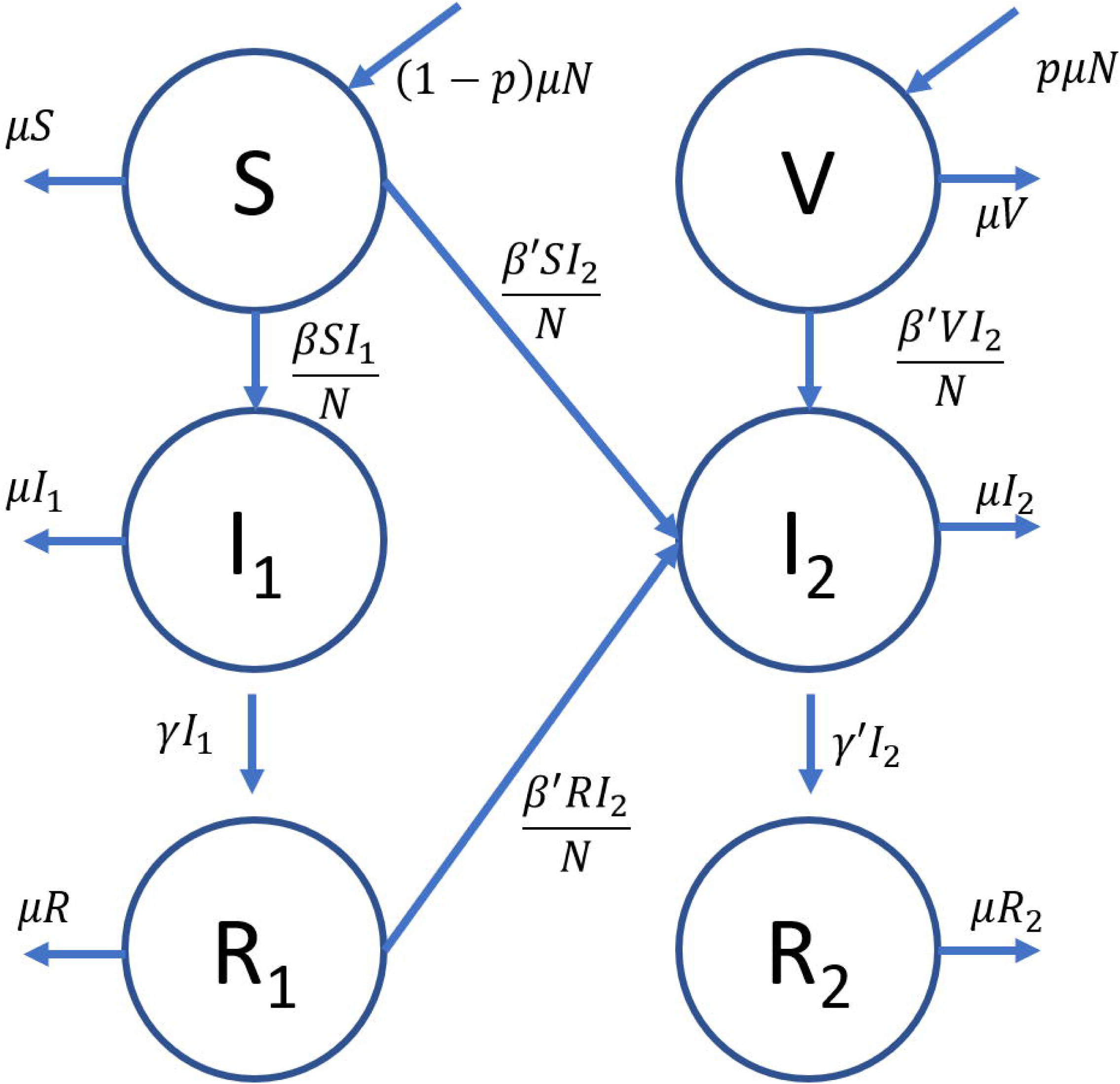
Compartment diagram for the emerging disease model. The transitions between compartments, together with the corresponding rates, are described by the arrows directed in and out of each compartment.

As in Chauhan et. al’s [34] work, the standard incidence was used to model infection of the susceptible individuals for the newer strain. Based on Eq 1 and the compartment diagram in Fig 2, the dynamics of the system can be expressed in terms of the following ordinary differential equations:

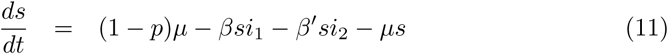

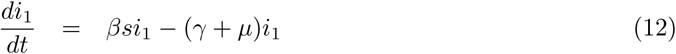

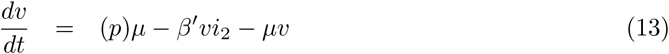

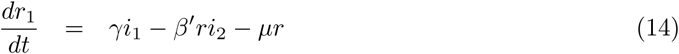

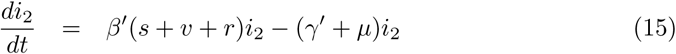

and *r*_2_ = 1 − *s* − *i*_1_ − *v* − *r*_1_ − *i*_2_. Similar to the simple SIR with vaccination scenario, the solution for the variables should follow the constraint

*s*(*t*), *i*_1_(*t*), *v*(*t*), *i*_2_(*t*), *r*_1_(*t*), *r*_2_(*t*) ≥ 0 for any time *t*.

To solve for the equilibrium points of the system, Eqs 11-15 should be equal to zero. Wolfram Mathematica [49] was used to obtain solutions for the system of equations, which are the following:

1. DFE:(*s,i*_1_,*v,r*_1_,*i*_2_) =(1−*p*,0,*p*,0,0)
2. Original strain equilibrium:

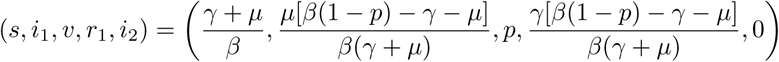
3. New strain equilibrium:

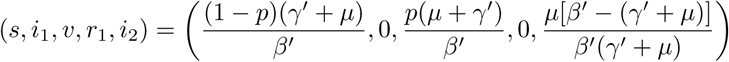
4. Endemic equilibrium: (*s, i*_1_, *v, r*_1_, *i*_2_) = (*s*^*^, *i*_1_^*^, *v*^*^, *r*_1_^*^, *i*_2_^*^)

The second equilibrium point corresponds to the scenario where only the original strain is present. Applying the constraint for the plausible solution, the original strain equilibrium exists if

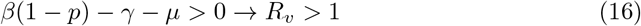

where *R*_*v*_ = *R*_0_(1 − *p*) = *β*(1 − *p*)*/*(*γ* + *μ*) is the reproduction number of the original strain for the SIR model with vaccination [34].

The third equilibrium corresponds to the scenario where only the new strain survives. For this equilibrium point to exist, the following condition should be satisfied:

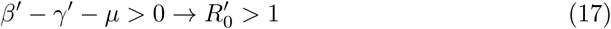

where 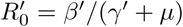 is the corresponding reproduction number of the newer strain if modeled using a standard SIR model.

Equilibrium point 4 is the endemic equilibrium where

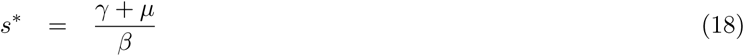

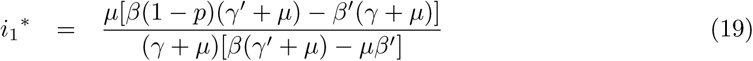

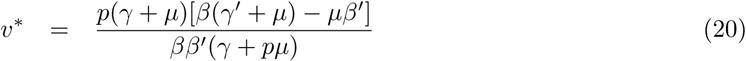

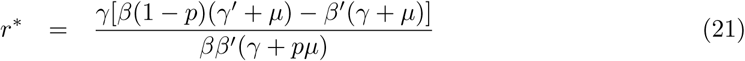

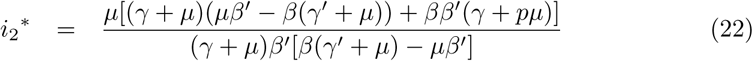

For the endemic equilibrium to exist, the following condition should be satisfied:

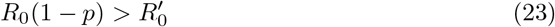

The next section will discuss the local stability of the four equilibrium points.

## Stability analysis and simulations

After solving for the equilibrium points, we need to determine the conditions in which these points are stable. These conditions dictate which equilibrium will describe the steady state behavior of the system. The local stability of the equilibrium points will be determined based on the eigenvalues of its Jacobian evaluated at a specific equilibrium point [17]. Let ***C***_**0**_ = (*C*_1_, *C*_2_, …)^*T*^ be the vector of the population number of each compartment. For a general compartment *C*_*i*_, the components of the Jacobian, *J*_*ij*_ can be obtained using the following equation:

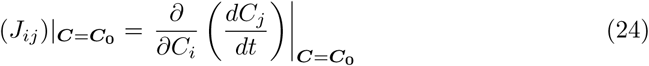

For our system, the Jacobian of the system can be obtained by applying Eq 25 to Eqs 11-15. For any equilibrium point, 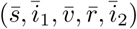 yields

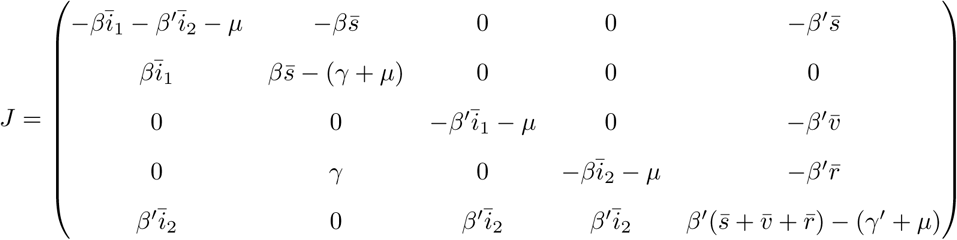

Local stability is attained when the eigenvalues of the Jacobian, ***λ***, are negative or have negative real parts. In other words, the solutions for ***λ*** such that det(***J*** − 𝟙*λ*) = 0 should be negative or have negative real parts if the solution is complex [50].

Simulations will then be used to check if the system approaches the equilibrium point. As described in the previous sections, the system starts as a one-strain SIR model with vaccination as discussed in the section “Modelling the emergence of the new strain” with the following values for the parameters: *μ* = 0.2 (birth/death rate) and *p* = 0.7 (vaccination rate). For this simulation, the time is discretized in terms of the average time between compartment interactions, i.e. the average time it takes for individuals to transition from one compartment to another. At time *t* = 0, we allow 1 % of the population to be infected by the original strain and the system is made to evolve in time using the values of infection coefficients *β* and removal rate *γ* that satisfies the respective requirements for the Reproduction number for each equilibrium point to exist‥ The new (mutated) strain was made to emerge at time point *t* = 100, when we expect the system to be in equilibrium. The new strain is introduced to the population by infecting 1% of the susceptible population with the newer strain. The evolution will then be dictated by the modified multi-strain SIR model developed in Section “Modelling the emergence of the new strain” using values for *β′* and *γ′* that satisfy the conditions for 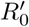 for each equilibrium point to exist.

### Disease free equilibrium (DFE)

The Jacobian for the DFE can be obtained by substituting the respective values to 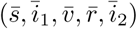 in the expression for the Jacobian. This yields:

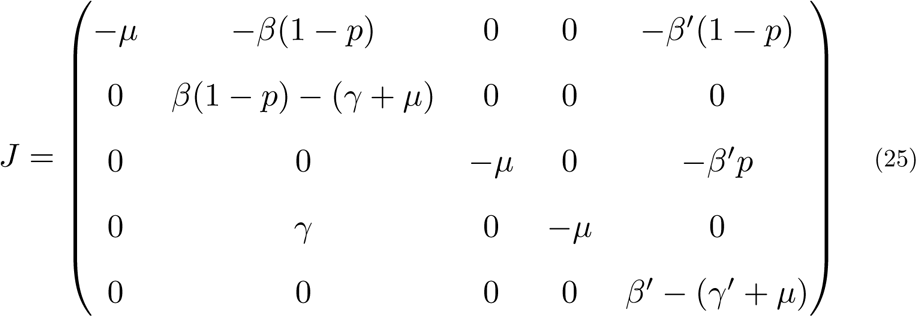

and the corresponding characteristic equation is

(*λ* − *μ*)^3^(*λ* − *β*(1 − *p*) + *γ* + *μ*)(*λ* − *β′* + *γ′* + *μ*) = 0. This means that the eigenvalues are *λ* = −*μ*, −*μ*, −*μ, β*(1 − *p*) − *γ* − *μ, β′* − *γ′* − *μ*. Recall that for the DFE to be locally asymptotically stable, all eigenvalues should have negative real parts. Hence, the following conditions should hold:

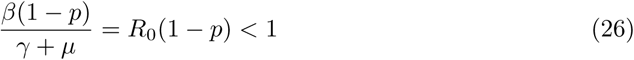

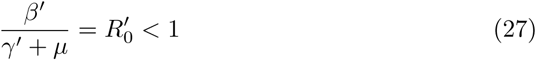

This is consistent with the local stability of the disease-free equilibrium for the regular standard incidence SIR and the SIR with vaccination models [34]. The conditions stated in Eqs 26 and 27 give us the threshold conditions of the transmission and removal coefficients for this model, which is related to the basic reproduction number of the model to be discussed in “Reproduction Number” section.

Eqs 26 and 27 also imply that if the system is in DFE before the emergence and the reproduction number of the emergent disease is less than that of the original disease, then the system will remain in DFE in the long run. Fig 3 shows the simulation of the DFE using parameters that satisfy Eqs 26 and 27. Note that the plot legends will be the same for the succeeding surveillance plots for the other equilibrium plots. The plot shows that the proportion of vaccinated individuals (*v*), denoted by the solid blue line, and the proportion of susceptible individuals (*s*), denoted by the solid black line, remained relatively constant at long times. Since the vaccination rate is set to be 0.7, we expect more individuals to be vaccinated than susceptible. Due to the emergence of the new strain at time point *t*=100, there appears to be a slight dip in *s* but it quickly stabilized to the disease free equilibrium.

**Fig 3.**
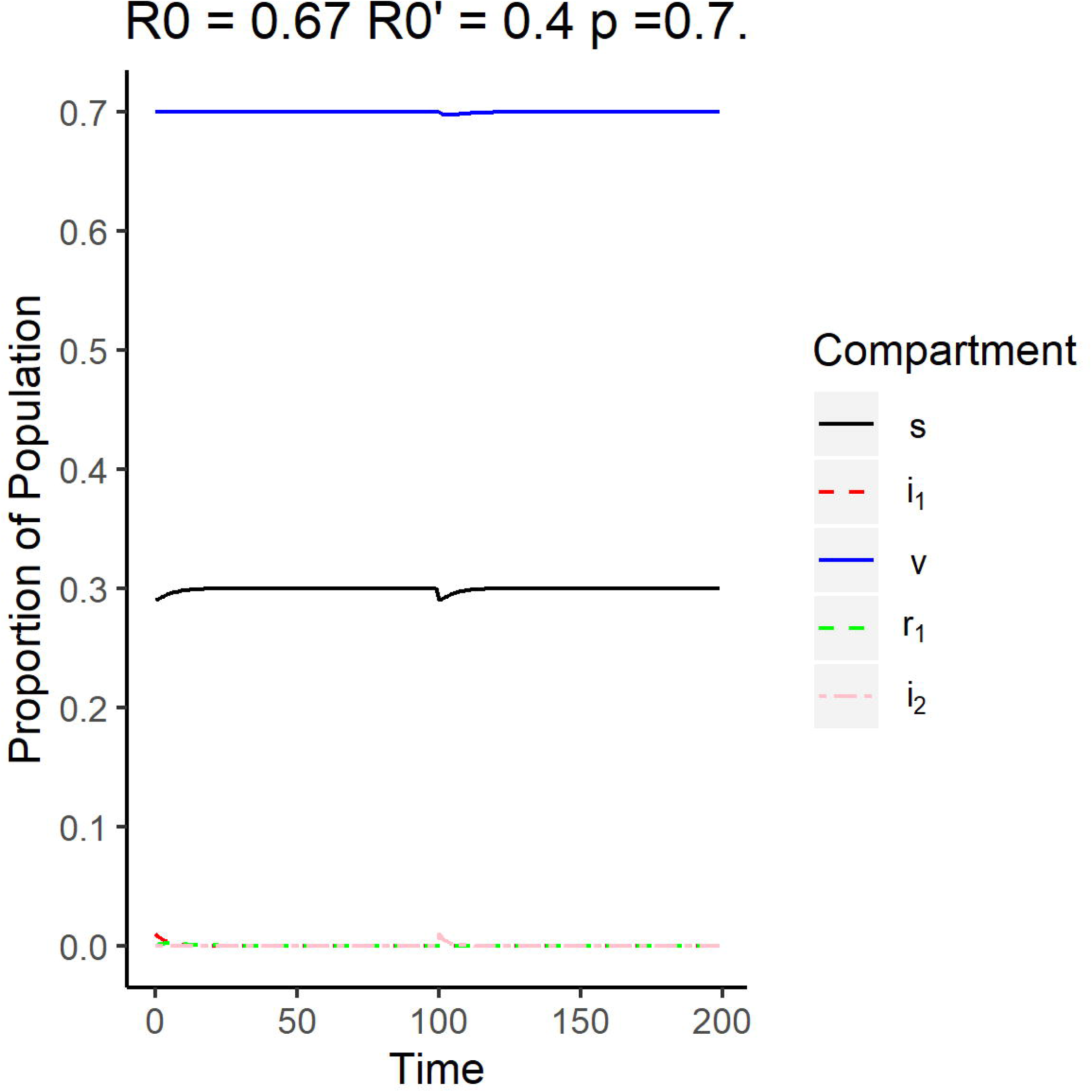
Surveillance data of the compartments for the disease free equilibrium. The reproduction number of the original strain is *R*_0_ = 0.67, while the reproduction number of the emergent strain is 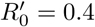. The vaccination rate used is 0.7.

### Disease 1 equilibrium (Original strain)

For this equilibrium scenario, *i*_1_ ≠ 0 and since Eq 12 is equal to zero then

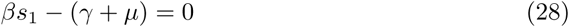

where *s*_1_ is the equilibrium value corresponding to the susceptible compartment for the Disease 1 equilibrium. We can calculate the resulting Jacobian for Equilibrium point 2 by substituting the corresponding values to the Jacobian equation given in Eq 25. The Jacobian is then given by,

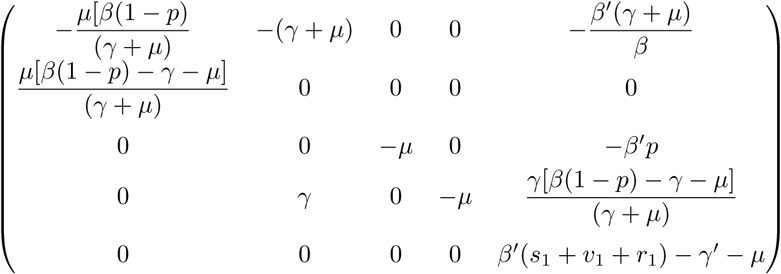

where *s*_1_, *v*_1_, *r*_1_ are the respective equilibrium values for the susceptible, vaccinated, and the initially recovered compartments for the Disease 1 equilibrium. The resulting eigenvalues are 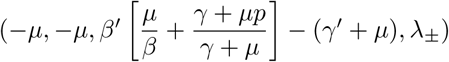, where

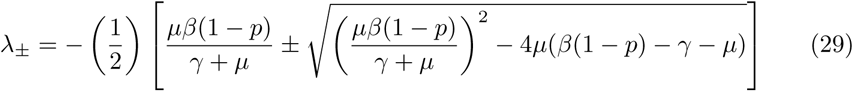

The discriminant of *λ*_±_ can dictate whether the eigenvalues will have a negative real part. If the discriminant is negative or zero, then the eigenvalues will be negative. If the discriminant is positive, recall that for the system to not go to the DFE, *R*_0_(1 − *p*) > 1.

This means that

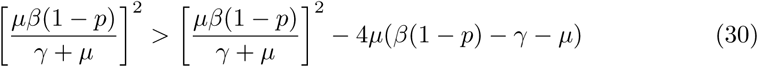

which ensures that *λ*_±_ is negative when *R*_0_(1 − *p*) > 1. When *R*_0_(1 − *p*) < 1, *λ*_±_ would have a positive real part which makes the equilibrium point unstable. This suggests that this equilibrium point will not be stable if the system was not already in endemic equilibrium with Disease 1.

For the third eigenvalue to be negative,

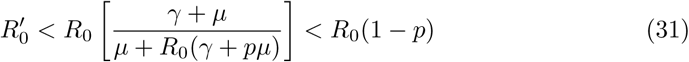

When these conditions are satisfied, then the Disease 1 equilibrium point is locally asymptotically stable. These conditions also imply that this equilibrium point can only be achieved if the system before the emergence is already in endemic equilibrium with Disease 1. Fig 4 shows the simulation of the system using parameters that satisfy Eq 31. Unlike the DFE case, the proportion of individuals infected by the original strain, denoted by the red dashed line, along with the recovered individuals, denoted by the green dashed line, increase up to their respective equilibrium values. This is accompanied by the decrease in susceptible individuals in the population.

**Fig 4.**
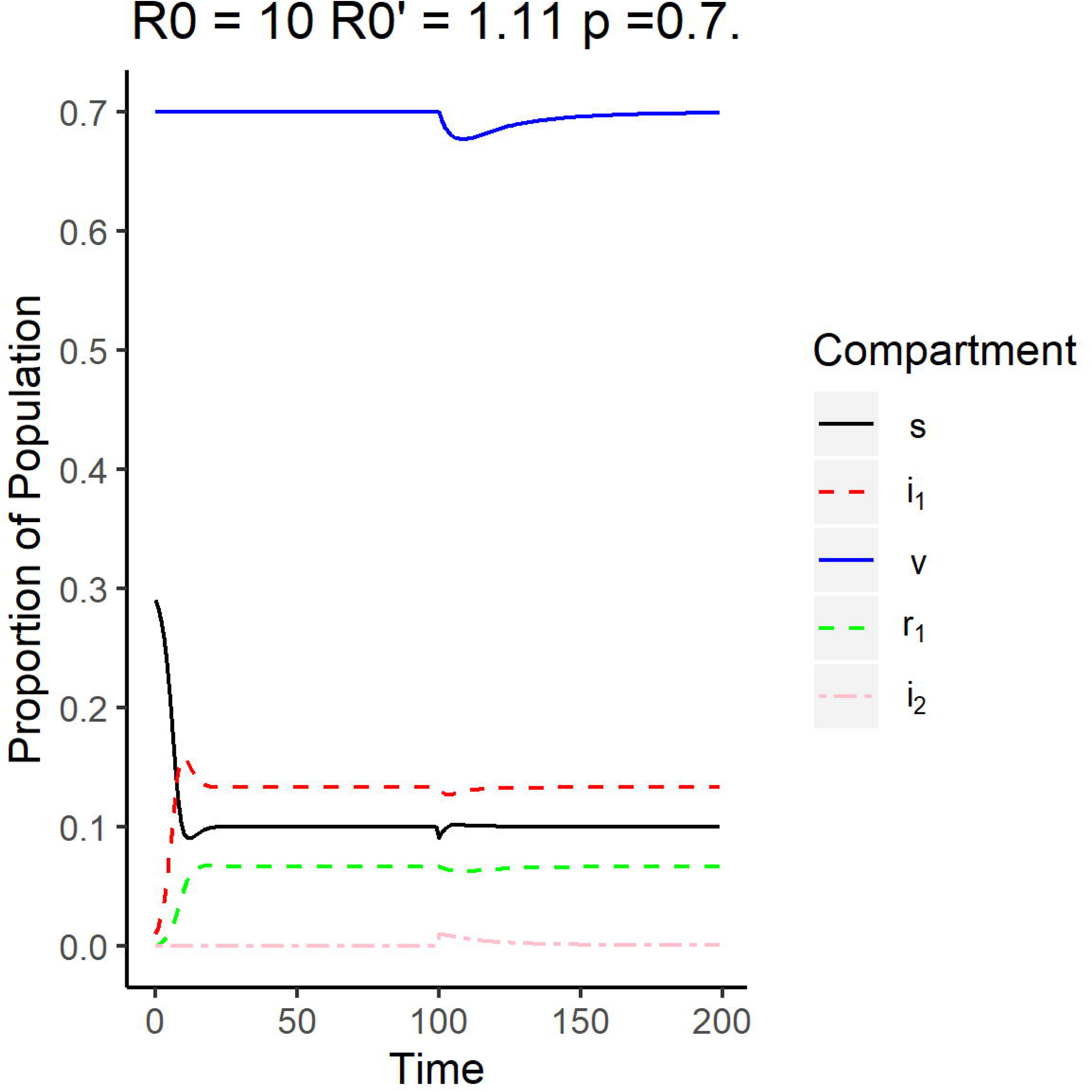
Surveillance data of the compartments for Disease 1 equilibrium. The reproduction number of the original strain is *R*_0_ = 10, while the reproduction number of the emergent strain is 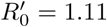. The vaccination rate used is 0.7.

At *t* = 100, the number of individuals infected by the newer strain, denoted by the pink dashed line, was shown to have a small spike, but quickly went to zero while the number of individuals infected by the original strain remained relatively unchanged. Note that the reproduction number of the emergent strain is 1.11 which is greater than one, meaning the emergent strain will be able to survive on its own in this population. This implies that the new strain is not strong enough to infect enough people to dominate over the original strain.

### Disease 2 equilibrium (Newer strain)

For this equilibrium scenario, *i*_2_ ≠ 0 and thus for Eq 15 to be zero, we have to have

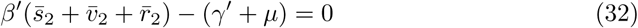

where 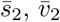, and 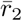 are the respective equilibrium values of the susceptible, vaccinated, and initially recovered compartments corresponding to the Disease 2 equilibrium. The resulting Jacobian, *J*, for the equilibrium where only the mutated disease exists is given by

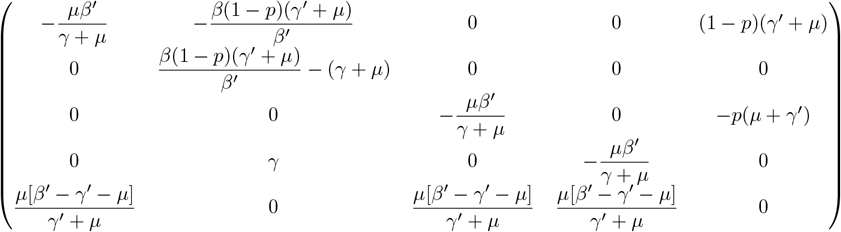

The corresponding characteristic equation is given by,

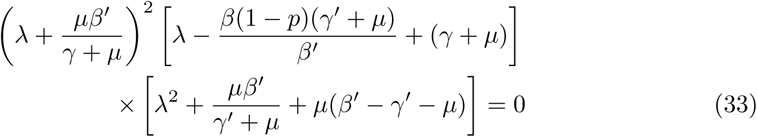

The eigenvalues are 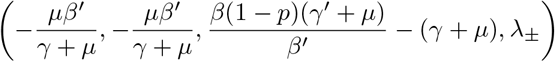
where

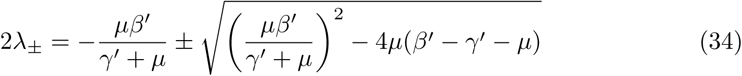

Similar to the *λ*_±_ in the Disease 1 equilibrium, the real part will be negative if the discriminant is negative. For the equilibrium point to exist, 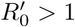, which means that when the discriminant is positive, the following inequality holds

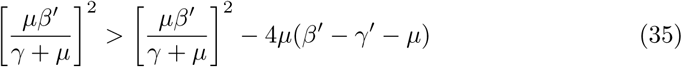

which means that both *λ*_±_ will be negative as long as 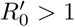. As for the remaining eigenvalue, it will be negative if

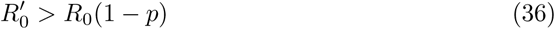

This indicates that the second disease will be locally stable if the mutated disease has a higher reproduction number compared to the original disease. Once this happens, the endemic equilibrium can not be achieved. Note that there are no conditions for the value of *R*_0_(1 − *p*), which means that this equilibrium point can occur whether the system was initially in DFE or endemic equilibrium with the original strain. Fig 5 shows the emergence from a system in DFE where *i*_1_ is zero before the emergence of the strain, which happens when 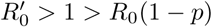. The proportion of vaccinated and the susceptible individuals remained constant before the emergence. Upon the emergence of the newer strain, the system behaves like a regular SIR model with a susceptible compartment comprising of the S, V, and R compartments as shown in Fig 5. The proportion of both susceptible and vaccinated individuals decreased drastically shortly after the emergence at *t* = 100, while the proportion of individuals infected by the emergent strain (*i*_2_), denoted by the pink dashed line, had an upward spike before settling into its equilibrium value. The original strain showed no sign of reemergence after it has settled into DFE, which is what is expected.

**Fig 5.**
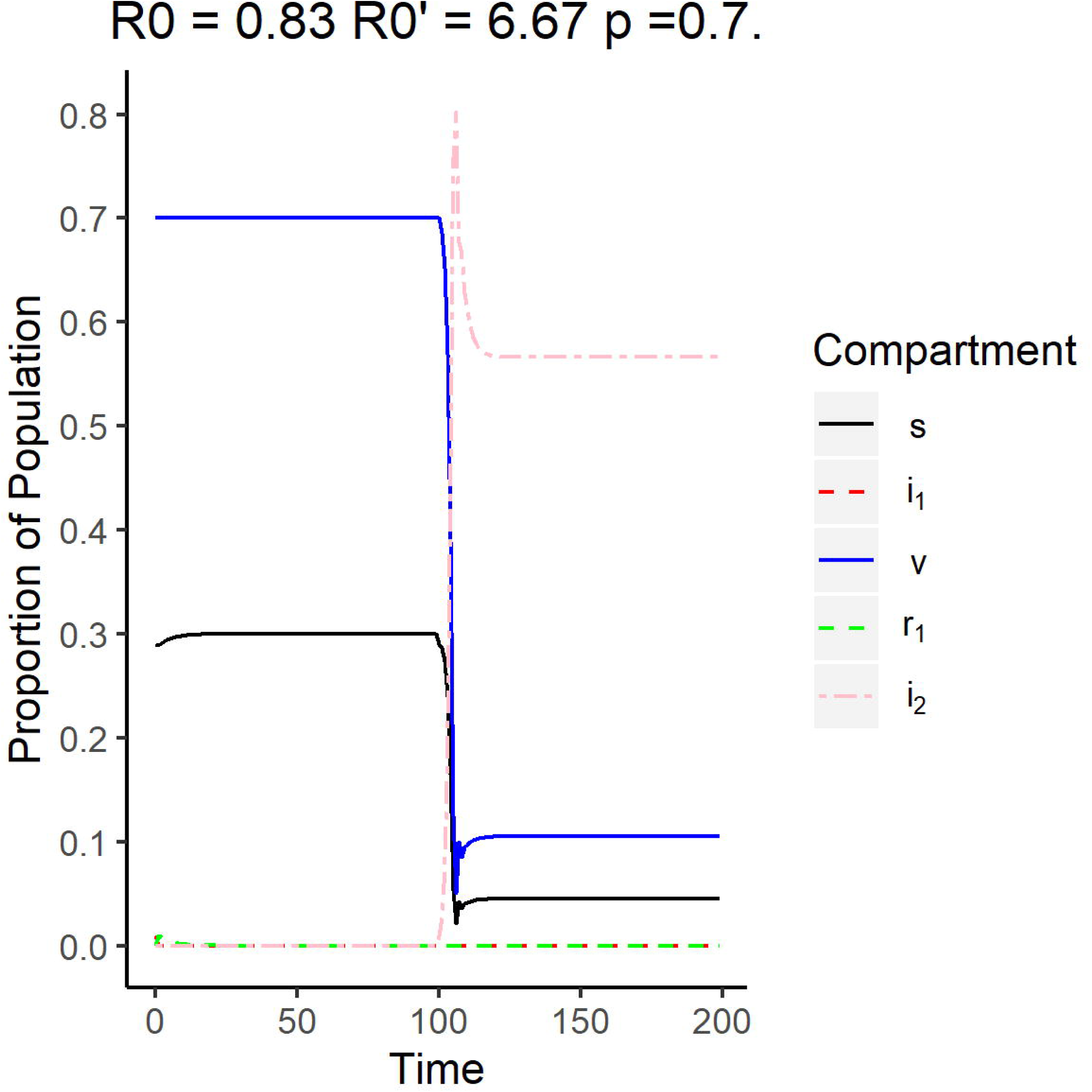
Surveillance data of the compartments for the new strain equilibrium where the system is originally in DFE. The reproduction number of the original strain is *R*_0_ = 0.83, while the reproduction number of the emergent strain is 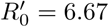. The vaccination rate used is 0.7.

Another possible case is when the system is initially in endemic equilibrium but the original strain dies because of the introduction of the new strain. Fig 6 shows that *i*_1_ is nonzero before time *t* = 100, which occurs when 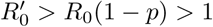. Upon emergence of the second strain, the proportion of the population infected by the original strain, *i*_1_, and the proportion of the individuals who recovered from the original strain, *r*_1_, decrease and go to zero asymptotically. The behavior of the second strain is similar to that in Fig 5. This implies that the new emergent strain is much more infectious than the older strain that the new strain infects more susceptible individuals compared to the original strain. This causes the original strain to die down at steady state.

**Fig 6.**
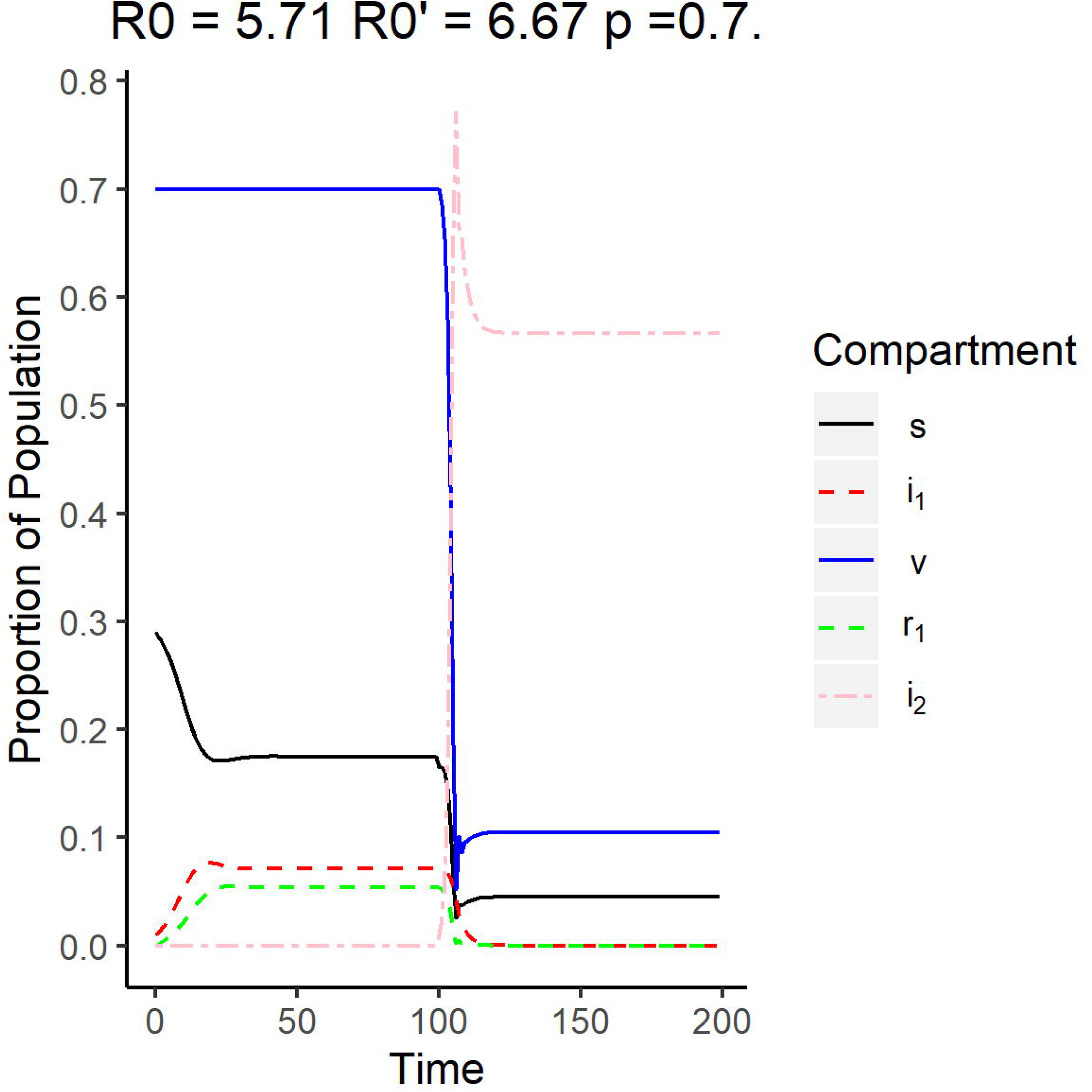
Surveillance data of the compartments for the new strain equilibrium where the system is originally in endemic equilibrium with Disease 1. The reproduction number of the original strain is *R*_0_ = 5.71, while the reproduction number of the emergent strain is 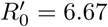. The vaccination rate used is 0.7.

### Endemic equilibrium

For the endemic equilibrium case, both *i*_1_ and *i*_2_ are nonzero and thus both Eq 28 and 32 hold. Since Eq 11 is zero,

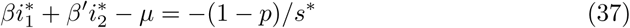

The resulting Jacobian for the endemic equilibrium case is given by

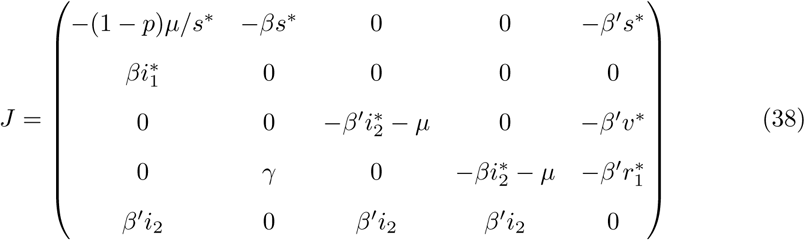

The characteristic equation is given by

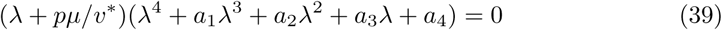

where

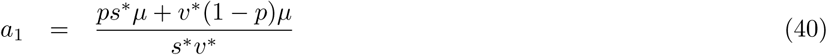

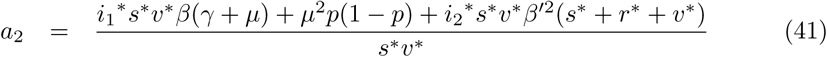

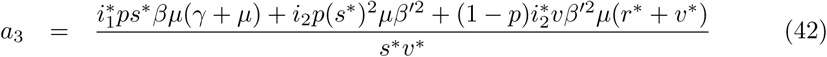

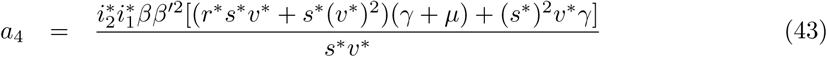

Recall that for the endemic equilibrium to be locally stable, all eigenvalues should have a negative real part. Eq 39 shows that one of the eigenvalues is *λ* = −*pμ/v*^*^ which is negative. For all the roots of the quartic term to have negative real parts, the Routh-Hurwitz criteria for stability should be applied [16, 17]. According to the Routh-Hurwitz criterion, a polynomial with degree 4 will have roots (*a*_1_, *a*_2_, *a*_3_, *a*_4_) that all have negative real parts when:

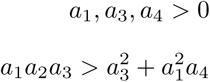

Since the values of the equilibrium points should be positive, all coefficients (*a*_1_, *a*_2_, *a*_3_, *a*_4_) are positive. Based on the stability of the first three equilibrium points and the existence criterion, the endemic equilibrium point is expected to be stable when

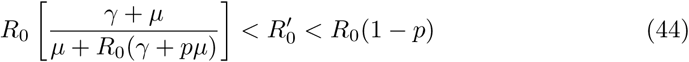

This implies that the endemic equilibrium for the two strains can only occur when the system is initially in endemic equilibrium for the original strain, which can explain why the condition is more restrictive than the one for the equilibrium with Disease 2. This highly restrictive criterion for endemic equilibrium might be a challenge for simulating stochastic data for strains that have a relatively low reproduction number and a high vaccination rate. A fluctuating value for the incidence rate might lead to either one of the single-strain equilibria.

Fig 7 shows that both *i*_1_ and *i*_2_ are asymptotically nonzero after the emergence of the new strain given that Eq 44 is satisfied.

**Fig 7.**
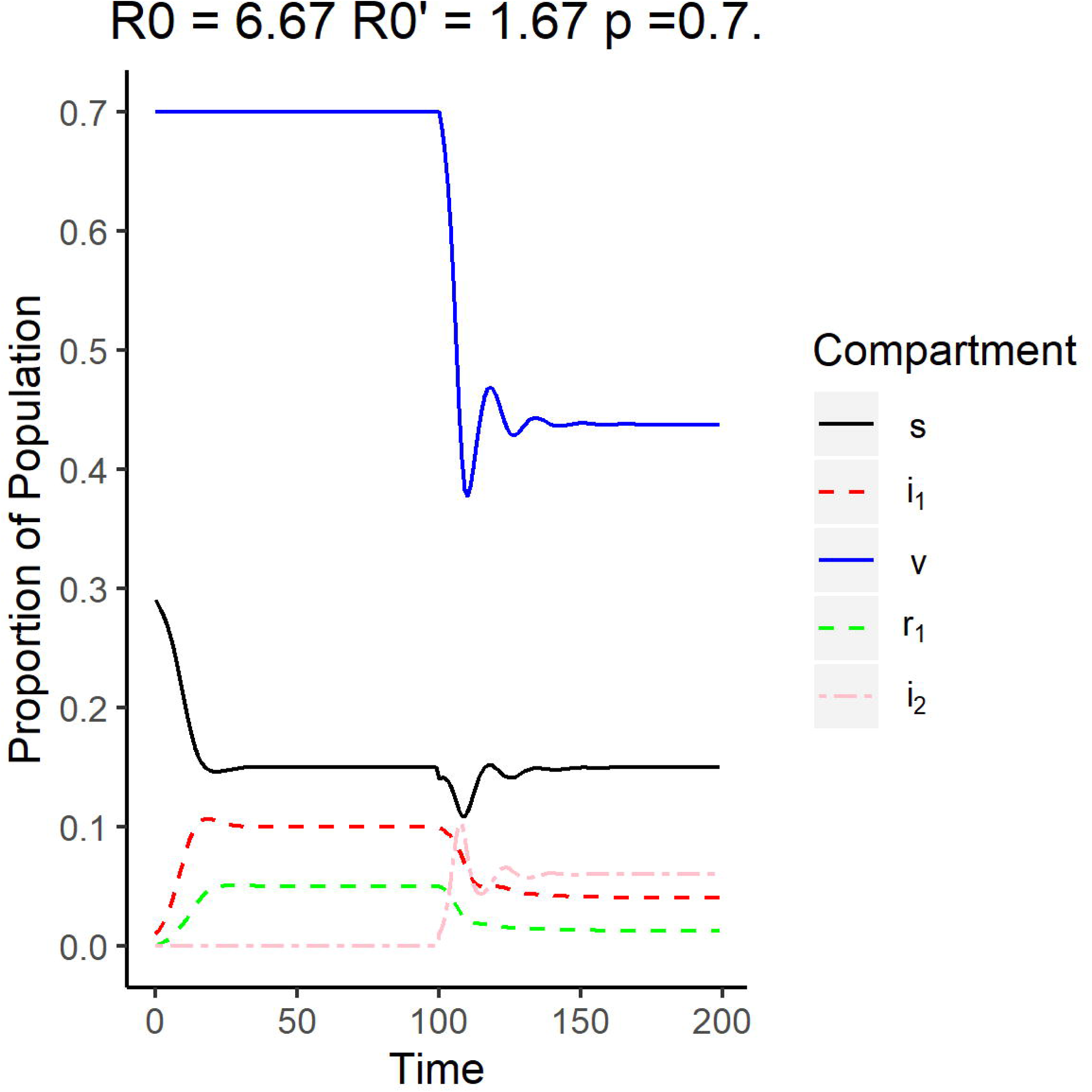
Surveillance data of the compartments for the endemic equilibrium. The reproduction number of the original strain is *R*_0_ = 6.67, while the reproduction number of the emergent strain is 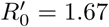. The vaccination rate used is 0.7.

As the endemic equilibrium between the two strains is reached, the proportion of the vaccinated individuals that are healthy decreased considerably compared to the proportion before the emergence while unvaccinated individuals return to the same proportion after the system has stabilized after the emergence of the new strain. This implies that the newer strain mostly survives on infecting the vaccinated and the initially recovered individuals. The proportion of individuals infected by the original strain is also observed to decrease upon reaching endemic equilibrium after emergence, which implies that some of the susceptible population get infected by the newer strain.

## Reproduction number [28]

One of the important parameters to be calculated for an epidemic model is the reproduction number, which quantifies how infectious a certain disease is. Formally, the reproduction number is defined as the expected number of secondary infections caused by a single infected individual for the whole duration that they are infectious. A value for the reproduction number that is greater than one indicates that the epidemic persists in the population, while a value less than one means that the disease will die out in the population [28, 29].

The reproduction number R_0_ of this epidemic model was calculated using the approach formulated by van den Diessche and Watmough [28]. This approach does not account for any measures taken to control the epidemic, but will give us an idea of the conditions needed for the disease to spread on its own.

### Next generation matrix

To obtain the reproduction number for this model, we need to solve for the next generation matrix. The next generation matrix describes the expected number of new infections that an infected individual produces from each susceptible compartment. Eqs 11 to 15 can be written in vector form as

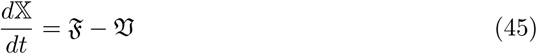

where 𝕏 = (*i*_1_, *i*_2_, *s, v, r*_1_)^*T*^, 𝔍_*i*_ corresponds to the vector that describes the rate of new infections in compartment *i*, and 𝔙_*i*_ is the vector that correspond to the transitions from compartment *i* to the other non-infected compartments such as *R*_1_ and *R*_2_ [51]. We define *F* and *V* such that for the disease free equilibrium 𝕏 _0_,

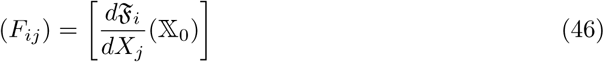

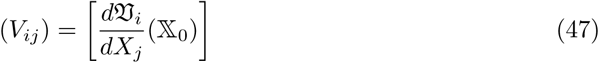

where (*i, j*) corresponds to the index of the infected compartments. The resulting matrices *F* and *V* have a dimension of *m* × *m*, where *m* is the number of infected compartments. *F*_*ij*_ describes the rate at which the infected individuals at compartment *j* contribute to the infection of compartment *i*, while *V*_*ij*_ corresponds to the rate at which the infected individuals are removed from the infected compartments. This means that *FV* ^−1^ is related to the rate at which individuals are infected by the disease within an average time span that an infected individual remains infected. For the system discussed in this paper, there are two infected compartments after the emergence of the new strain: *I*_1_ and *I*_2_. Therefore, *m* = 2 and 1 ≤ *i, j* ≤ *m*.

The DFE was calculated to be given by (1 − *p*, 0, *p*, 0, 0). Based on the transition equations (Eqs 11 to 15), F and V are given by

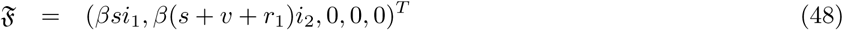

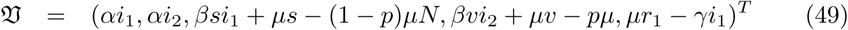

where *α* = *γ* + *μ*. Note that *m* = 2, so the corresponding *F* and *V* matrices are then

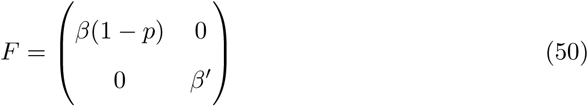

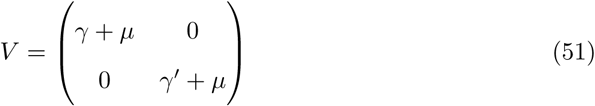

The reproduction number is obtained by taking the maximum eigenvalue of the next generation matrix *FV* ^−1^. The next generation matrix is the product of the rate of infection (*F*) and the average time that an individual remains infected (*V* ^−1^). The next generation matrix is given by,

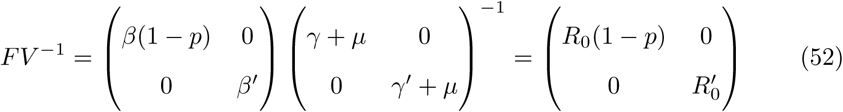

where *F* is the matrix that describes the infection rates for the two infections at the DFE, and *V* ^−1^ describes the average time an infected individual stays infectious. It is easy to see that the eigenvalues of the next generation matrix are *R*_0_(1 − *p*) and 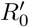. This means that the reproduction number R_0_ for this system is given by the larger of the two. Formally,

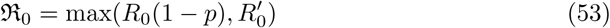

Note that the resulting threshold equation for the system is R_0_ less than one, which means that the system will only approach the disease free equilibrium when Eq 53 is less than one. For an outbreak to occur, at least one of these two strains should be able to persist in the population on its own, that is, to have an individual reproduction number greater than one. This is consistent with Eqs 26 and 27 which give us the condition of the stability of the DFE.

## Discussion and recommended next steps

We started modeling the emergence of a new strain by adding and modifying compartments to the existing SIR model with vaccination. The emergent strain was assumed to be unaffected by the existing vaccine designed for the original strain in the population. After establishing the possible transitions between compartments, the system was found to have four equilibrium points: the disease free equilibrium, the existing strain equilibrium, the emergent strain equilibrium, and the endemic equilibrium. Upon examining the conditions for existence and local stability, the disease-free equilibrium was determined to be locally stable when both *R*_*v*_ and 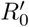 are less than one. This is consistent with the reproduction number for this multi-strain model. The existing strain equilibrium surprisingly did not impose that 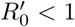, which is the condition for the DFE of a normal SIR model for a single-strain without immunity. This implies that if the original strain is much more contagious than the emergent strain, the emergent strain would still die out eventually regardless of the fact that it would persist in the population if it was on its own. On a similar note, the local stability condition for the emergent equilibrium condition also does not impose that *R*_*v*_ < 1. This means that the state of the system prior to the emergence of the new strain does not matter as long as the emergent strain is more contagious than the existing strain. This leaves a highly restrictive condition for endemic equilibrium to exist: the emergent strain must be able to survive by itself and the original strain must be contagious enough to infect enough people, which is given by Eq 44. Eq 44 also implies that the two-strain endemic equilibrium will only be locally asymptotically stable if the original strain is endemic to the population upon the emergence of the newer strain. These restrictions for the stability of endemic equilibrium would highly affect simulation studies about emergent strains especially when stochasticity is added to the model. Knowing the stability conditions for a multi-strain SIR model without cross-immunity will also be able to give us insights about when a newer strain emerges while the original strain still exists. This is common for the flu virus which changes every season and the vaccines lose their efficacy after a new mutated strain emerges. This result might also be relevant when a highly contagious strain like the COVID-19 virus, a viral strain that does not exhibit cross-immunity with the influenza virus, emerges in a population. Our results show that COVID-19 will be able to co-exist with the flu in an endemic equilibrium under certain circumstances, which might become a problem in terms of prioritizing patients in health care centers especially since the two diseases show similar symptoms [52]. Our research also suggests that if the spread of the emergent virus is not contained, we can expect the COVID-19 virus to dominate over the existing influenza virus, which can highly influence the development of protocol in receiving patients with flu-like symptoms and allocating resources in health care centers. In summary, the modified multi-strain SIR model of an emerging disease that affects both susceptible and previously immune individuals was studied. The local stability of the equilibrium points as well as the reproduction number for the model were calculated. Based on the results, we found that the original and the emergent strain can coexist in an endemic equilibrium if the emergent strain has a lower reproduction number than the original strain and that the system should already be in endemic equilibrium with the original disease before the emergence. The requirement for the endemic equilibrium to exist is quite strict especially for low values of *R*_0_ and high values of *p*, which presents a challenge in simulating surveillance data with stochastic incidence rates.

This modified SIR model can be improved further by using time-dependent infection rates to account for the seasonality of viruses. Exploring the effect of vaccination on other epidemic models such as epidemics with animal vectors and epidemics with latency and treatment compartments can also be studied in the future.

## Data Availability

This study was mostly theoretical and used simulated data from R for illustration. The code and the data underlying the results presented in the study are available from.

## Acknowledgments

We would like to acknowledge Dr. Yu Jin and Dr. Clay Cressler for their input on the mathematical derivation and applications of the model.

## Notes

### Competing Interest Statement

The authors have declared no competing interest.

### Funding Statement

The authors received no specific funding for this work.

